# Vanderbilt Integrated Community TMS for Opioid Recovery (VICTORY): Study protocol for a randomized, controlled trial of non-invasive brain stimulation to reduce craving in people with opioid use disorder

**DOI:** 10.64898/2026.08.18.26360768

**Authors:** Kathryn Biernacki, Jillian Connolly, Libby Tunison, Kristopher A. Kast, Simon Vandekar, Benson King, Tashley Aouina, Beth Black, Rachel Craig, Jason Ferrell, Christopher A. Grimes, Lauren Horowitz, Michael Levin, Mariah Smith, Linda Sok, Amanda von Horn, Kyle York, Steven Somers, Jonathan Becker, Michelle Cochran, Heather Burrell Ward

**Affiliations:** Department of Psychiatry & Behavioral Sciences, Vanderbilt University Medical Center, Nashville TN 37232; Department of Biostatistics, Vanderbilt University Medical Center, Nashville, TN 37232; Synaptic PSYCH, Brentwood TN 37027

**Keywords:** Opioid use disorder, transcranial magnetic stimulation, craving, intervention

## Abstract

**Background:** Individuals receiving buprenorphine treatment for opioid use disorder (OUD) remain at high risk for treatment discontinuation and return to opioid use. Transcranial magnetic stimulation (TMS) has shown efficacy in reducing craving and substance use in other substance use disorders, but its application in OUD remains limited and the neural mechanism underlying its therapeutic effects is poorly understood. Determining the feasibility and generalizability of TMS in patients receiving buprenorphine - the most commonly prescribed medication for OUD - is therefore critical. This protocol aims to address these issues in a clinical trial of weekly TMS sessions for OUD.

**Methods:** We will enroll up to 120 individuals with OUD taking buprenorphine in a randomized, single-blind, sham-controlled trial of left dorsolateral prefrontal cortex (DLPFC)-targeted intermittent theta burst stimulation (iTBS). Participants will receive active or sham iTBS weekly (2 sessions of 1800 pulses each applied once per week x 8 weeks, 16 sessions total) with pre- and post-iTBS assessments (10, 12, 20 weeks) of craving, opioid use, and treatment retention. A subset of individuals will undergo optional pre- and post-iTBS neuroimaging. The study will be conducted at an academic medical center and a private outpatient TMS clinic.

**Aims:** Our primary aim is to determine whether 16 sessions of active iTBS applied to the left DLPFC results in reduced craving and opioid use, and higher treatment retention, relative to sham. In a secondary aim, we will also examine whether iTBS-related changes in craving are associated with changes in functional connectivity between the left DLPFC and both the dorsal striatum and anterior cingulate cortex.

**Discussion:** By evaluating the feasibility and efficacy of a weekly TMS protocol that aligns with routine care and focuses on patients maintained on buprenorphine, this study addresses key limitations of prior TMS research in OUD. Furthermore, the inclusion of neuroimaging will help characterize the neural mechanisms underlying TMS-related changes in craving.

**Trial registration:** This clinical trial is registered at ClinicalTrials.Gov; ID NCT07457489; date of registration: 03/02/2026.

## Introduction

Opioid use disorder (OUD) affects over 16 million people each year and remains the leading cause of fatal overdoses worldwide (World Health Organization, 2023). Gold-standard medications for the treatment of OUD, such as buprenorphine, the most widely prescribed medication for OUD (Shulman, Wai, & Nunes, 2019), safely reduce mortality and drug use (Degenhardt et al., 2011; Volkow, Jones, Einstein, & Wargo, 2019). However, even with buprenorphine treatment, relapse and treatment discontinuation rates remain high for people with OUD; the three-month period after treatment initiation presents the highest risk for return to use (Lee et al., 2018; Opheim et al., 2021). Within 6 months of starting buprenorphine, only 50% of patients will continue treatment, and 91% will return to use (Weiss et al., 2011). Together, these data highlight the need for adjunctive treatments that address the drivers of return to use and treatment discontinuation.

Noninvasive brain stimulation has been explored as a treatment for substance use disorders (SUDs), including OUD. Transcranial magnetic stimulation (TMS) is one form of noninvasive brain stimulation that uses electromagnetic pulses to modulate cortical excitability and induce neuroplastic changes (Hallett, 2007; O’Shea & Walsh, 2007). The U.S. Food and Drug Administration (FDA) has cleared TMS for the treatment of depression, obsessive-compulsive disorder and for smoking cessation (Cohen, Bikson, Badran, & George, 2022) with clinical protocols typically administering stimulation over the dorsolateral prefrontal cortex (DLPFC; Kan et al., 2023). Within these applications, intermittent theta burst stimulation (iTBS), a specific type of patterned TMS, has demonstrated significant clinical utility. iTBS is thought to increase synaptic long-term potentiation by enhancing cortical excitability (Blumberger et al., 2018; Suppa et al., 2016). Crucially, reviews and meta-analyses across multiple SUDs have shown that TMS (including iTBS) is effective at reducing craving and substance use (Coles, Kozak, & George, 2018; Hone-Blanchet, Ciraulo, Pascual-Leone, & Fecteau, 2015; Mehta et al., 2024). It is hypothesized that TMS exerts therapeutic effects by targeting the neural circuits that contribute to drug craving and seeking (Hanlon, Dowdle, Lench, & Ramos, 2020).

Despite these promising findings, research examining TMS in OUD remains limited, with reviews of noninvasive brain stimulation in OUD identifying only a handful of studies (Blyth et al., 2025; Ward, Mosquera, Suzuki, & Mariano, 2020; Young et al., 2020). Existing studies of TMS for OUD primarily focus on individuals using heroin and only one study has assessed the impact of accelerated deep TMS in people receiving buprenorphine for OUD (Guldas, Tumkaya, & Yucens, 2025). To date, only four studies have investigated TMS for OUD in North America – where the use of the highly potent synthetic opioid, fentanyl, has been steadily increasing and presents a higher risk of overdose than heroin (Substance Abuse and Mental Health Services Administration, 2025). Two of these studies have used single-session TMS protocols to modulate craving or cognition (Biernacki et al., 2025; Steele, 2023). One recently published study evaluated an accelerated TMS protocol in which 4 sessions of iTBS were delivered in a single day to a small sample of patients with OUD who were stable on their maintenance medications (Ballard et al., 2026). The authors reported reduced attentional bias to opioid cues, however this did not reach statistical significance. The third study implemented a repeated-sessions clinical protocol with TMS administered daily over multiple weeks (Tang et al., 2025). However, of 87 individuals screened for participation, only 6 were randomized, and 4 completed the study. Accordingly, no statistical analyses were conducted on this small sample, thus limiting the evaluation of clinical efficacy and treatment outcomes. Notably, none of these studies evaluated individuals taking buprenorphine, despite it being the most commonly prescribed medication for OUD in North America (Shulman et al., 2019). As such, the following questions remain about the use of TMS for OUD:

1. Is TMS for OUD effective, and if so, how does it exert its effects? Only one study of TMS for OUD has included neuroimaging (Steele, 2023), despite neuroimaging being critical for helping to characterize the neural circuit changes that occur with TMS treatment.
2. Is a weekly dosing schedule of TMS for OUD a feasible and acceptable adjunctive treatment? A previous study (Tang et al., 2025) testing therapeutic TMS protocols (i.e., daily sessions of treatment over multiple weeks) struggled with recruitment and retention, questioning the feasibility and acceptability of this intervention for this population.
3. Can we provide TMS for populations with OUD beyond urban academic medical centers? Expanding access to TMS has been a challenge for the field more broadly, besides the treatment of SUDs. While clinical trials for TMS are typically conducted in academic medical centers, more widespread clinical application would ultimately require delivery of TMS for SUDs in private outpatient clinics nationwide.

To address these questions, we designed a multi-site randomized controlled trial of TMS for OUD among individuals receiving buprenorphine treatment. We will implement a novel weekly stimulation protocol designed to improve treatment efficiency and better align TMS delivery with routine weekly clinic visits. The trial will be conducted across both an academic medical center and private practice clinics throughout Tennessee to help address the question of clinical applicability beyond academic medical settings. Our central hypothesis is that DLPFC-targeted iTBS will significantly reduce both opioid craving and opioid use. In secondary analyses, we will use neuroimaging (MRI) to assess whether a change in functional connectivity between the DLPFC and both the dorsal striatum and anterior cingulate cortex (ACC) is associated with reductions in craving. Ultimately, this work aims to establish a safe and effective adjunctive therapy to directly mitigate the severe craving and high relapse rates that limit current OUD treatments.

## Methods

### Study Design

The study is a randomized, single-blind, sham controlled clinical trial of iTBS to reduce opioid craving in people diagnosed with OUD who are currently receiving treatment with buprenorphine. The study consists of 12 study visits. Participants will also have the option to elect to participate in optional MRI scans before and after the intervention. At the first visit (Consent), participants will complete informed consent, which will be obtained by the researcher conducting the study. After completing consent, participants will undergo additional screening procedures to ensure safety to receive TMS (and safety to receive MRI if the participant elects to complete these scans) and will complete questionnaires (listed in Figure 1) about substance use and psychiatric symptoms. Enrolled participants will then undergo 8 weekly sessions of left DLPFC-targeted iTBS. Prior to the first TMS visit, individuals will be randomized to receive active or sham iTBS. At each TMS visit, participants will complete questionnaires about their substance use and psychiatric symptoms and provide a sample for urine toxicology. After the 8-week TMS intervention, participants will return for follow-up visits at weeks 10, 12, and 20. At each follow-up visit, participants will complete questionnaires about their substance use and psychiatric symptoms and provide a sample for urine toxicology. If a participant elects to undergo the optional MRI visits, they will undergo 2 MRI scans 1) before the first TMS visit; and 2) after the week 8 TMS visit. During both MRI scans, participants will complete a resting state fMRI and perform a task assessing cue reactivity in the scanner. At these sessions, participants will also complete tasks outside the scanner assessing inhibitory control and reward processing. See Figures 2 and 3 for a visualization of the study design and Figure 1 for a list of assessments conducted on each study visit. This protocol was developed with input from multiple certified peer recovery specialists (T.A., C.G., K.Y.), who are coauthors on this manuscript and have lived experience with substance use.

**Figure 1.**
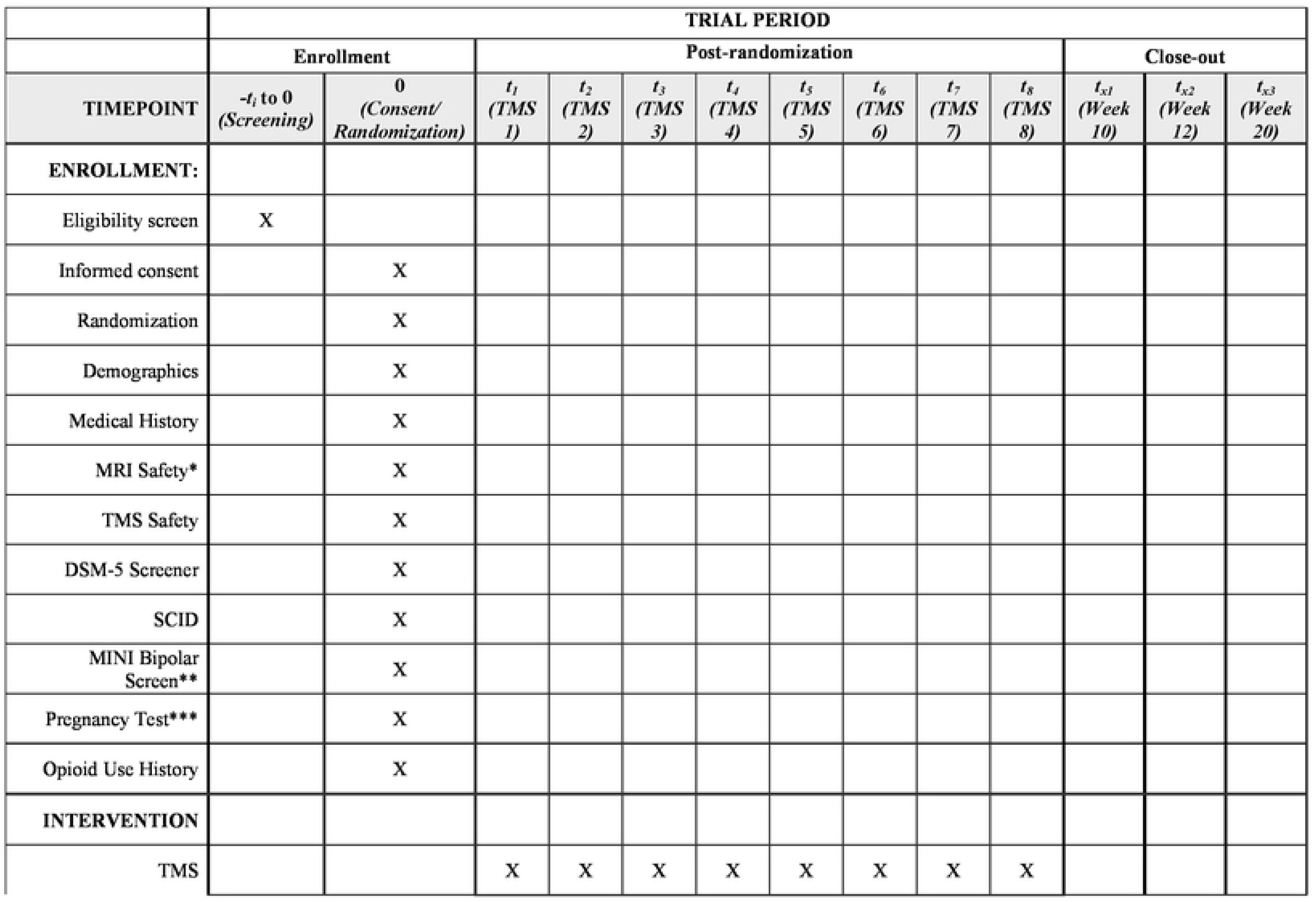
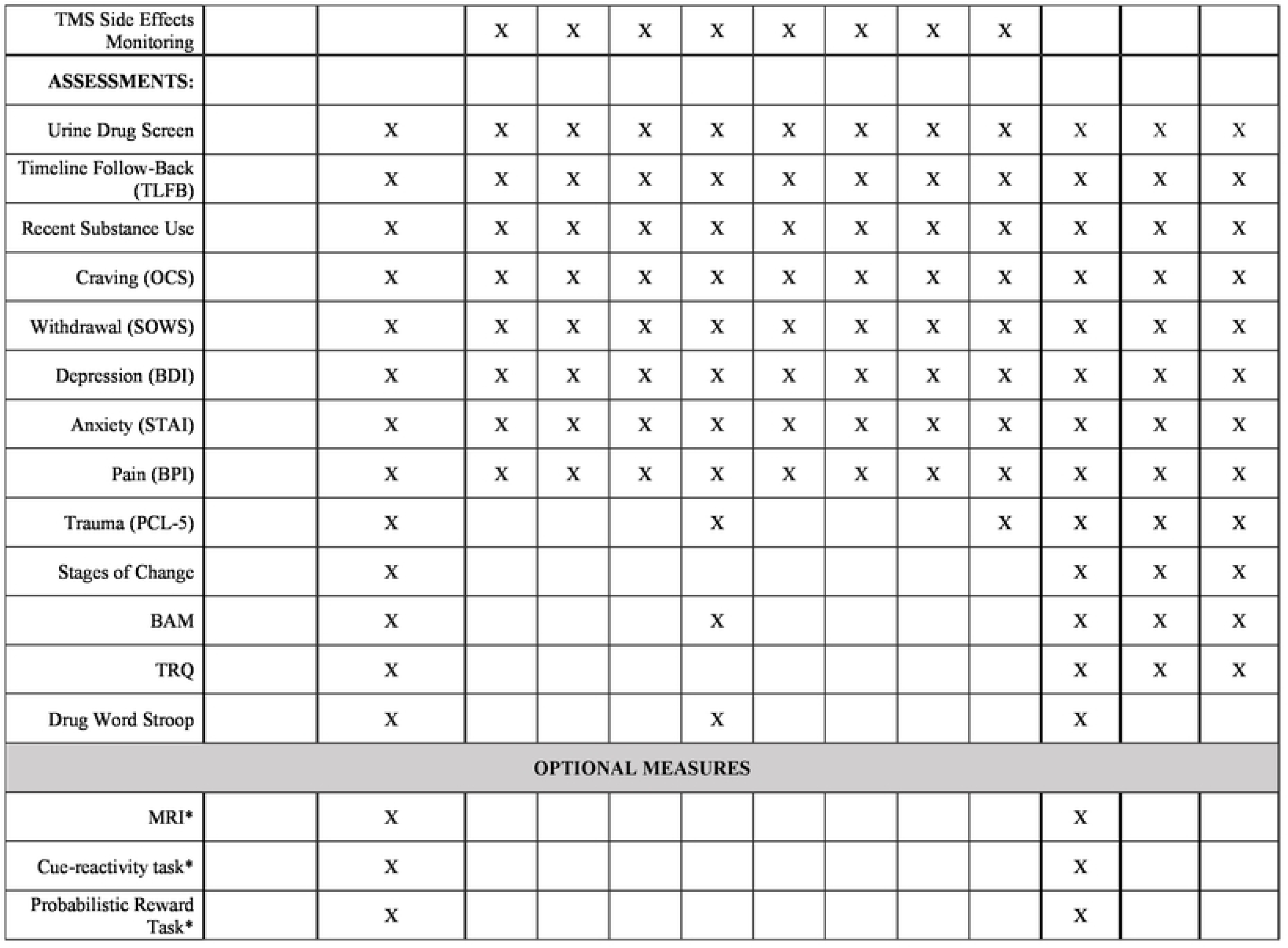
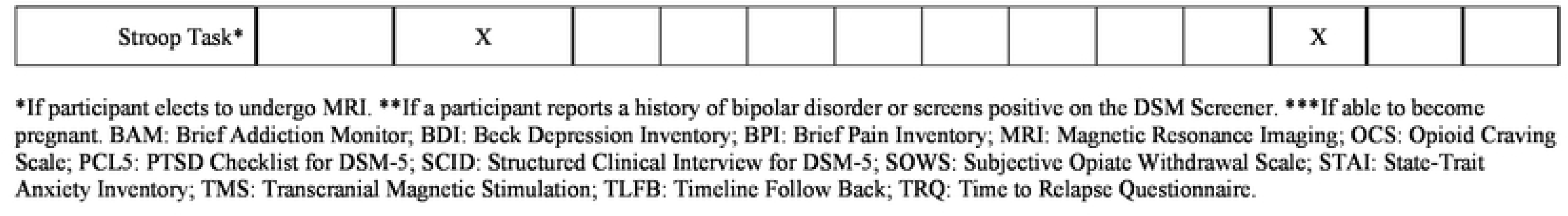
Participant timeline: Schedule of enrollment, interventions, and assessments. *If participant elects to undergo MRI. **If a participant reports a history of bipolar disorder or screens positive on the DSM Screener. ***If able to become pregnant. BAM: Brief Addiction Monitor; BDI: Beck Depression Inventory; BPI: Brief Pain Inventory; MRI: Magnetic Resonance Imaging; OCS: Opioid Craving Scale; PCL5: PTSD Checklist for DSM-5; SCID: Structured Clinical Interview for DSM-5; SOWS: Subjective Opiate Withdrawal Scale; STAI: State-Trait Anxiety Inventory; TMS: Transcranial Magnetic Stimulation; TLFB: Timeline Follow Back; TRQ: Time to Relapse Questionnaire.

**Figure 2.**
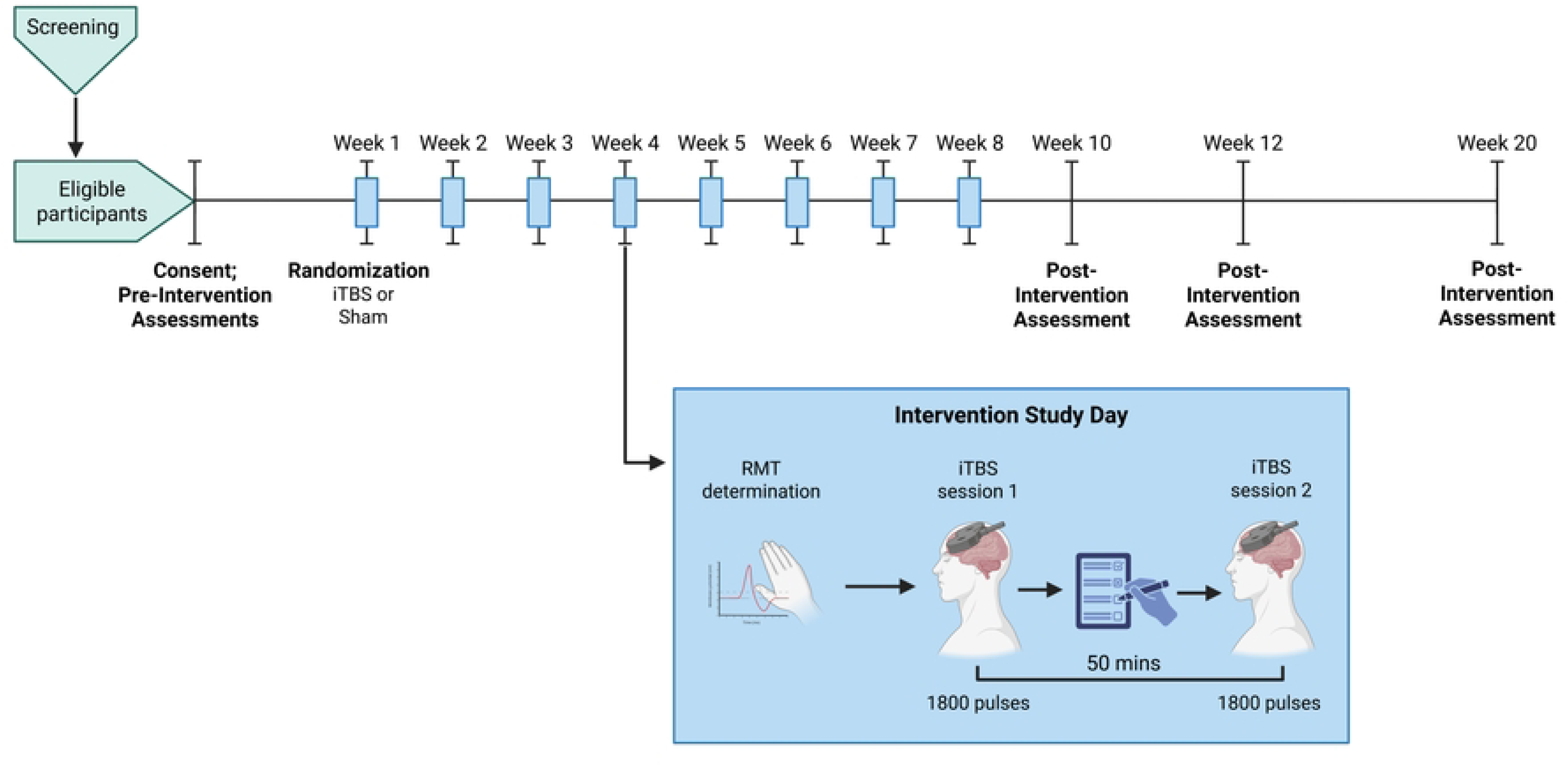
Study Protocol Diagram. After consent and randomization to receive active or sham stimulation, participants will receive iTBS weekly (2 sessions once per week x 8 weeks) with pre- and post-intervention assessments of craving, opioid use, and treatment retention. RMT: resting motor threshold.

**Figure 3.**
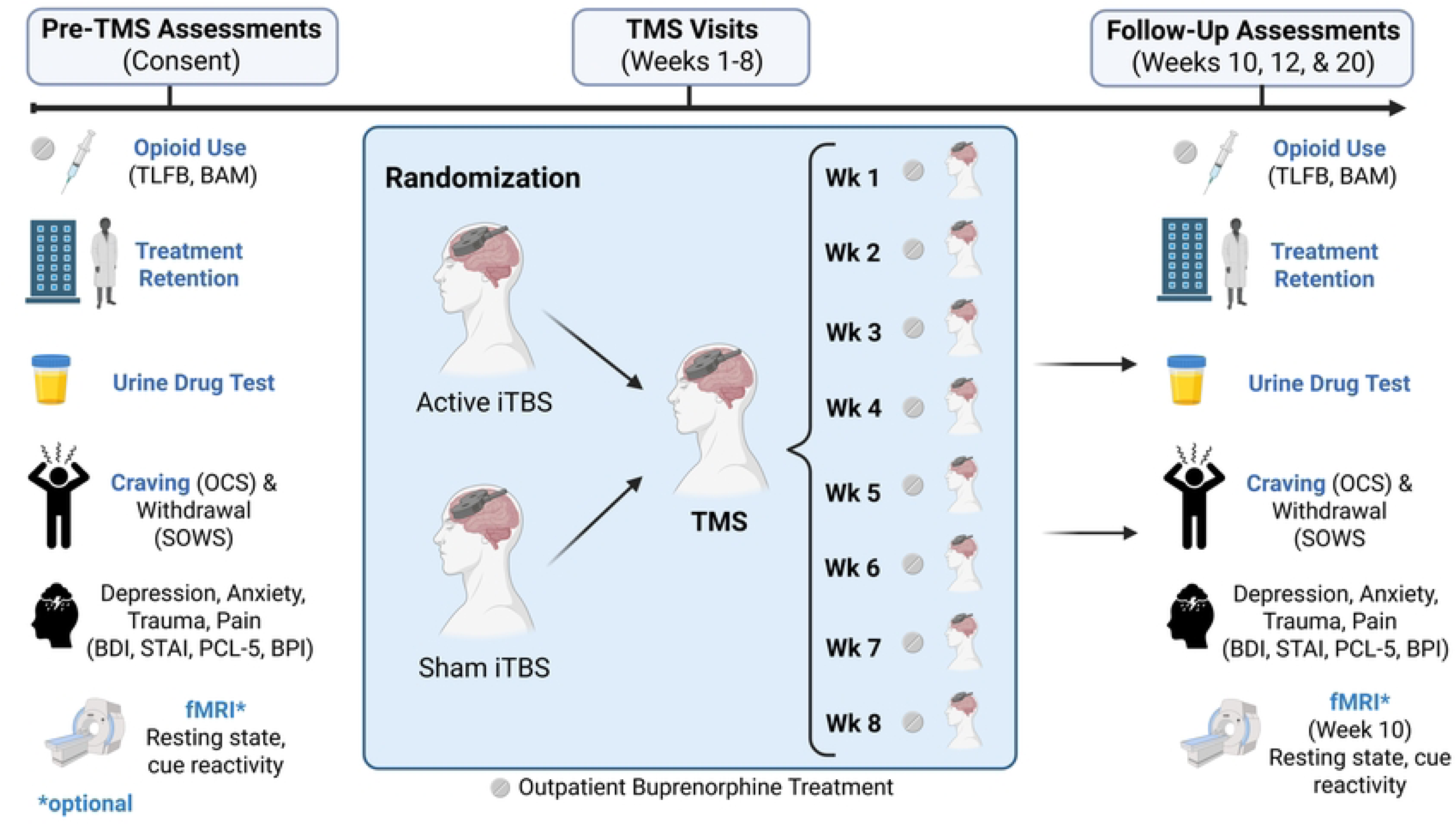
Diagram of schedule of assessments. Participants will complete assessments of opioid use, treatment engagement and retention, drug use, craving and other psychiatric assessments before and after receiving TMS treatment. A subset of participants will also complete an optional neuroimaging session at baseline and post-TMS where measures of craving and cognition will be collected.

### Study Approval and Registration

This protocol is approved by the Vanderbilt University Medical Center (VUMC) Institutional Review Board (IRB) and Advarra IRB (VUMC Protocol ID: 251293; Advarra Protocol ID: Pro00094328). Modifications to the protocol (at both sites) will be communicated to the study teams by electronic notification (e.g., email). The protocol has also been registered at ClinicalTrials.gov (ID: NCT07457489). Results from this trial will be reported in ClinicalTrials.gov and will be published in academic journals. Results may also be disseminated at academic and medical conferences.

### Study Setting

The study will include two enrolling sites, one of which is a medical academic institution (VUMC) and the other a private psychiatric clinic (SynapticPSYCH). Both sites have experience conducting research using TMS methods. Research ethics approval for the academic institution is covered by an institutional IRB. Research ethics approval for the clinic site is covered by an external central IRB. Protocol amendments will be submitted to the institutional IRB and external central IRB, which will send electronic notifications or changes required directly to the participating sites.

### Inclusion and Exclusion Criteria

#### Inclusion Criteria

- Age between 18-65 years
- Diagnosis of OUD according to DSM-5 criteria and confirmed by SCID (First, Williams, Karg, & Spitzer, 2015)
- Currently prescribed buprenorphine for opioid use disorder
- Meets either of the following criteria: (1) reports opioid craving of 2 or greater on a 0-10 scale, or (2) has returned to opioid use at least once within the past 12 months
- Must be able to read, speak and understand English
- Must be judged by study staff to be capable of completing the study procedures
- Participants will be in stable outpatient psychiatric treatment and psychiatrically stable.

#### Exclusion Criteria

- DSM-5 intellectual disability
- Substance use disorder (other than opioid, nicotine, or cannabis) within the past 3 months
- Current, active suicidal ideation with intent or plan
- Positive urine drug screen for illicit substance use that can increase seizure risk (cocaine, benzodiazepines, amphetamine, methamphetamine)
- History of psychosis in the past 3 months or diagnosis of a primary psychotic disorder
- Any history of a progressive or genetic neurologic disorder (e.g. Parkinson’s disease, multiple sclerosis, tuberous sclerosis, Alzheimer’s Disease) or acquired neurological disease (e.g. stroke, traumatic brain injury, tumor), including intracranial lesions
- History of head trauma resulting in any loss of consciousness (>15 minutes) or neurological sequelae
- Current history of poorly controlled headaches including chronic medication for migraine prevention
- History of fainting spells of unknown or undetermined etiology that might constitute seizures
- History of seizures, diagnosis of epilepsy, or immediate (1st degree relative) family history epilepsy with the exception of a single seizure of benign etiology (e.g. febrile seizures) in the judgment of a board-certified neurologist
- Chronic uncontrolled medical conditions that may cause a medical emergency in case of a provoked seizure (e.g., cardiac malformation, cardiac dysrhythmia, asthma, etc.)
- Any metal in the brain or skull (excluding dental fillings) or elsewhere in the body unless cleared by the responsible covering physician (e.g. MRI compatible joint replacement)
- Any devices such as pacemaker, medication pump, nerve stimulator, TENS unit, ventriculo-peritoneal shunt unless cleared by the responsible covering physician
- All female participants of child-bearing age will be required to have a pregnancy test; any participant who is pregnant or planning to become pregnant will not be enrolled in the study
- Medications will be reviewed by the responsible covering physician and a decision about inclusion will be made based on the participant’s past medical history, drug dose, history of recent medication changes or duration of treatment, and use of CNS active drugs. The published TMS guidelines review of medications to be considered with TMS will be taken into consideration given their described effects on cortical excitability measures
- Participants who, in the investigator’s opinion, might not be suitable for the study or would be unable to tolerate the study visit

### Sample Size

We plan to recruit up to 60 participants at each site (VUMC and SynpaticPSYCH) to obtain a completion sample of n = 50 at each site for a total of n = 100. With 80% power, a type 1 error rate of 5%, and 100 subjects across all sites, we can detect effect sizes larger than 0.40. Based on these calculations, our proposed total sample of n = 100 will provide adequate power to detect changes in craving and opioid use.

### Recruitment and Screening

Chart review and team meetings will be used to identify outpatients at the academic medical institution and at the private clinic who meet criteria for the proposed study diagnoses. The research team will speak with the patient’s psychiatrist and that individual will inform the research team whether they believe the patient can comprehend the study and procedures. If they deem the patient can provide informed consent and understand the study, the patient will be approached by someone familiar to them (e.g. a member of their care team) to see if they are willing to discuss participation in this study. If the patient assents, their name will be given to a research staff member who will contact them to schedule a study session. Patients may be pre-screened by phone. Research personnel involved with recruitment will discuss the study with the patient. If the participant meets criteria for the study, they may be scheduled for participation in the study. Research participants may be recruited through inpatient, partial hospital, intensive outpatient, or other clinics.

### Randomization and Blinding Procedures

Randomization will be stratified by site, with independent allocation sequences generated within each site. Within each stratum, treatment assignments will be generated using a permuted block design to maintain balance between groups throughout enrollment. The study biostatistician (SV) will generate the randomization sequence. As this is a single-blind study, study personnel responsible for administering TMS will have access to the allocation sequence, maintained on a central electronic document, and will not be blinded to treatment assignment. Participants will remain blinded to the intervention they receive throughout the duration of the study. Sham (placebo) stimulation will be delivered by flipping the coil 180 degrees so that the procedure mimics the look and sound of active stimulation but deliver no stimulation to the participant. Participants will only be un-blinded if an adverse reaction to active TMS occurs and the participant can no longer remain enrolled in the study. We will make numerous attempts to retain participants in the study with phone calls, emails, and text messages.

### Intervention

#### iTBS Equipment

The study will use the MagVenture MagPro X100 with MagOption (VUMC) or MagPro R30 (SynapticPSYCH) system equipped with a Cool-B70 butterfly coil to determine motor threshold and to administer the iTBS intervention. Active stimulation will be delivered with the coil in the active orientation. Sham stimulation will be delivered by flipping the coil 180 degrees so that this condition will look and sound like active stimulation but deliver no stimulation to the participant. TMS will be administered by Bachelors-level trained technicians under the supervision of a board-certified psychiatrist at both sites.

#### iTBS Treatment Dose and Frequency

Participants will receive 2 sessions of iTBS per study visit. iTBS will be delivered at 100% of resting motor threshold, which will be determined before the first iTBS of each study visit. Resting motor threshold (RMT) will be obtained by following recommendations from the International Federation of Clinical Neurophysiology (Groppa et al., 2012), which is defined as the lowest stimulus intensity required to elicit a visible motor twitch of the abductor pollicis brevis in 5 out of 10 consecutive trials. The iTBS intervention will use FDA-cleared parameters for depression (Cole et al., 2022; Cole et al., 2020). On each study day, iTBS will be applied over the left DLPFC twice, with each application consisting of 1800 pulses (60 cycles of ten 50-Hz triplets delivered in 2-second trains (5 Hz) with an 8-second intertrain interval). This protocol will occur twice per day (3600 pulses per day), separated by a 50-minute interval, which has been shown to be a safe and effective spacing interval for iTBS (Cole et al., 2022; Cole et al., 2020). This schedule will thus deliver a total of 28,800 pulses over 8 weeks (16 sessions).

### Monitoring

An independent safety monitor (ISM) will review this study for safety and adherence to the study protocol. The ISM will meet with the study PI every 6 months to assure safety of research participants, regulatory compliance, and data integrity. The ISM is distinctly separate from the role of the PI. A report will be generated after every ISM meeting and communicated with the IRB if needed. The ISM will review any seizure in order to assess what new precautions may be needed for subsequent subjects. A single occurrence of one of the following will trigger stopping the study, immediate ISM assessment, and FDA notification: seizure, status epilepticus, attempted or completed suicide. All participants will also be monitored throughout the study for any adverse reactions in relation to the TMS using the Adverse Reaction Form for TMS. Adverse reactions will be reviewed by a physician daily prior to TMS and then following TMS. The TMS sessions will be discontinued if the psychiatrist evaluates this side effect to be a worsening related to TMS application and the patient’s treating psychiatrist will be contacted if clinically indicated.

### Study Status and Timeline

Participant recruitment commenced on June 1^st^ 2026 at both study sites and remains ongoing as of August 2026. Recruitment is projected to be completed in June 2028. Final data collection is expected to conclude in November 2028, and study findings are anticipated to be available in December 2028.

### Outcomes and Assessments Primary Outcome

The primary outcome is to determine the effects of active left DLPFC targeted iTBS compared to sham on craving and opioid use in 100 people with OUD. Specifically, this study seeks to provide evidence that L DLPFC-targeted iTBS leads to reduced craving and opioid use in individuals with OUD.

Participants’ craving and drug use will be assessed prior to receiving iTBS, during iTBS intervention and following the end of iTBS (10, 12 and 20 weeks). Craving will be measured using the Visual Analogue Scale (VAS) on the Opioid Craving Scale (OCS). Opioid use will be measured as the number of days of use in the preceding week using the Timeline Follow back (TLFB).

### Secondary Outcome

In a secondary analysis, we will also determine if left DLPFC functional connectivity change is associated with change in craving on the OCS (n=50). If a participant elects to undergo the optional MRI visits, they will receive structural and resting-state functional magnetic resonance imaging (rsfMRI) scans. The rsfMRI scans will be used will be used to determine if active TMS affects left DLPFC resting-state functional connectivity more than sham and if change in functional connectivity between the left DLPFC and dorsal striatum and anterior cingulate cortex (ACC) is associated with change in craving score on the OCS.

### Statistical Analysis Plan

#### Primary outcome analysis

A change in craving score (Δcraving) on the OCS will be calculated between pre-TMS (week 1 timepoint) and post-TMS (week 10 timepoint) and a linear model will determine whether Δcraving significantly differs between active and sham conditions (controlling for age, sex, and pre-TMS craving). Change in number of days of use on the TLFB (Δuse) will be calculated between pre-TMS (week 1 timepoint) and post-TMS (week 10 timepoint) and a model will determine whether Δuse significantly differs between active and sham conditions (controlling for age, sex, and pre-TMS days of use). In post hoc analyses, we will investigate the effects of buprenorphine dose in response to TMS.

#### Secondary outcome analysis

As a secondary aim, we will investigate effects of TMS on left DLPFC resting-state functional connectivity. We will perform analyses to determine: 1) if there is an effect of TMS intervention (active vs. sham) on DLPFC-striatum connectivity and DLPFC-ACC connectivity; and 2) if change in DLPFC functional connectivity (post-pre) is associated with change in craving measured using the Opioid Craving Scale (n=50).

We hypothesize that active DLPFC-targeted iTBS will increase left DLPFC resting-state functional connectivity to the anterior cingulate cortex and striatum. Left DLPFC-striatum connectivity will be calculated by extracting the BOLD time courses from a seed placed in the left DLPFC and masks of the striatum and anterior cingulate cortex (ACC), calculating the Pearson correlation coefficient between the left DLPFC and these regions, then Fisher’s z-transforming the correlation estimate to generate metrics of DLPFC-striatum connectivity and DLPFC-ACC connectivity. We will fit a linear model using Δfunctional connectivity (post-pre) as the outcome and TMS intervention (active vs. sham) as the predictor, controlling for age, sex, and pre-TMS connectivity.

We also hypothesize that increased left DLPFC resting-state functional connectivity will be associated with reduced craving on the OCS. To test this, we will fit a linear model using Δcraving (post-pre) as the outcome and Δfunctional connectivity as the predictor, controlling for pre-TMS craving, pre-TMS functional connectivity, age, and sex.

We will also perform brainwide seed to voxel analyses of left DLPFC resting-state functional connectivity to determine alternative regions where active TMS changed connectivity more than sham and to identify alternative regions where ΔDLPFC functional connectivity was associated with Δcraving.

#### Data retention and missing data

We will collect data from all participants who attend visits. If missingness in outcome or covariate values are greater than 10%, we will perform multiple imputation by chained equations using 100 resampled datasets for the primary outcome analyses and secondary analyses that do not use full image outcome variables (Azur, Stuart, Frangakis, & Leaf, 2011; White, Royston, & Wood, 2011). We will compare results with and without multiple imputation as supplementary analyses. For full image analyses, we will use complete-case analysis. If a participant deviates from the protocol, we will perform sensitivity analyses using data collected per-protocol and including those who deviated from the protocol in separate analyses.

## Discussion

We describe a protocol for a multisite trial of TMS for OUD for individuals taking buprenorphine using a novel treatment schedule that optimizes for feasibility and efficiency. This protocol will evaluate the effectiveness of a therapeutic TMS intervention for reducing craving and opioid use in individuals receiving treatment for OUD. This protocol will extend prior work by administering TMS on a novel weekly schedule that more closely reflects typical clinical practice, and will focus on participants maintained on buprenorphine, the most commonly prescribed medication for OUD. In addition, the protocol will include neuroimaging to assess changes in neural circuitry associated with TMS intervention, which may help clarify potential mechanisms underlying treatment effects.

A key strength of this protocol is its focus on improving feasibility and generalizability of TMS for OUD - directly addressing limitations of prior clinical protocols. For example, a study conducted in China demonstrated TMS-related reductions in craving among individuals with OUD (Liu et al., 2020); however, participants were inpatients, were not receiving pharmacological treatment, and were exclusively using heroin. These conditions differ substantially from those in the United States, where most individuals receiving treatment for OUD receive outpatient treatment with medications such as buprenorphine and are required to attend regular clinic visits. In contrast, the only study to implement a therapeutic TMS protocol in North America reported significant challenges with recruitment and retention, requiring participants to attend sessions five days per week for four weeks (Tang et al., 2025). As noted by the authors, low retention likely reflects the complex nature of OUD, in which a range of social, medical, and personal factors can affect treatment engagement (Sahlem et al., 2024; Tang et al., 2025), and more broadly suggests that intensive daily TMS schedules may not be feasible in this population. These observations thus highlight the need to develop protocols that are better aligned with patient needs and real-world care contexts (Ward, Blyth, & Kast, 2025). Indeed, prior work suggests that the spacing of TMS treatments (e.g., daily vs weekly) is less critical than the total number of treatments or pulses delivered for providing therapeutic benefit (Fitzgerald et al., 2020; Galletly, Gill, Clarke, Burton, & Fitzgerald, 2012). Notably, participants enrolled in this protocol will receive a higher dose of TMS (i.e., more pulses) than the FDA-approved standard iTBS protocol (in which iTBS is delivered daily) while simultaneously fitting the treatment course into the cadence of weekly clinical visits for buprenorphine prescriptions. Additionally, while accelerated TMS protocols have shown promise in treating psychiatric conditions (Cole et al., 2022; Cole et al., 2020), the effect of spacing treatment is still untested. Thus, our approach of aligning TMS sessions with routine clinical visits may reduce participant burden without compromising dosage and may ultimately improve treatment adherence.

An additional strength of this protocol is the inclusion of a non-academic clinical site for data collection, which may enhance generalizability to real-world settings. Much of the existing TMS research has been conducted in academic medical centers. While these settings provide important infrastructure, they may not reflect the environments in which most patients receive care. As TMS becomes more widely available in private psychiatric clinics, evaluating protocols in these settings is increasingly important. By including a study site at a private psychiatric clinic, this protocol extends assessment to a more representative care setting, which may further enhance the feasibility and clinical applicability of the protocol beyond its clinical benefits.

Reducing craving, and in turn opioid use, is the primary clinical target of this protocol. Craving is a central feature of OUD (Kleykamp, Weiss, & Strain, 2019) and is the strongest predictor of drug use and relapse for people with SUD (Kakko et al., 2019; Lueptow, Shashkova, Miller, Evans, & Cahill, 2020; Vafaie & Kober, 2022). Craving is described as the subjective experience of wanting to use a drug (Tiffany & Wray, 2012) and exposure to cues that induce craving can more than triple the odds of drug use (Vafaie & Kober, 2022). Importantly, treatment with buprenorphine only partially attenuates craving, which can lead to a return to opioid use (Shulman et al., 2019). Consistent with this, previous work has directly linked craving to subsequent opioid use after buprenorphine treatment initiation (Tsui, Anderson, Strong, & Stein, 2014). Given the ubiquity of patient reports of craving and its hinderance to recovery, there has thus been a push to directly address craving in the treatment of OUD (Kleykamp, De Santis, et al., 2019; Kleykamp, Weiss, et al., 2019). Accordingly, the current TMS protocol may represent a complementary intervention to target this key driver of continued use and return to use, with the potential to improve clinical outcomes.

While improving outcomes is the first step toward implementing TMS in OUD populations, it is equally critical to understand the mechanisms underlying its therapeutic effects (Steele, 2021). To address this, we will incorporate neuroimaging to assess changes in neural circuitry associated with TMS intervention. Substantial evidence indicates that individuals with substance use disorders exhibit dysfunction in fronto-striatal circuits that increase drive for drug-seeking and reduce behavioral control (Feil et al., 2010; Goldstein & Volkow, 2002, 2011). Specifically, dysfunctional connectivity between the striatum and DLPFC is associated with increased drug use and impulsivity, dependence severity and increased cue-induced craving (Dugré, Orban, & Potvin, 2023; Gerchen, Rentsch, Kirsch, Kiefer, & Kirsch, 2019; Hu, Salmeron, Gu, Stein, & Yang, 2015; Yuan et al., 2017; Zhang et al., 2025). Importantly, a key mechanism by which TMS is thought to exert its therapeutic effects is via dopaminergic modulation of the striatum through stimulation of the DLPFC (Diana, 2011). Work has demonstrated that DLPFC stimulation promotes dopamine release in the striatum (Cho & Strafella, 2009; Strafella, Paus, Barrett, & Dagher, 2001) which may then counteract dopamine hypoactivity that is common in substance use disorders (Koob & Volkow, 2016; Nutt, Lingford-Hughes, Erritzoe, & Stokes, 2015). Together, these findings suggest that targeting DLPFC-striatal connectivity may serve as an effective intervention to reduce craving and return to use in individuals with OUD and highlight the importance of using neuroimaging to evaluate whether this TMS intervention engages these circuits and leads to clinically meaningful changes in craving.

While this study has several strengths, some limitations of the protocol highlight important unanswered questions in the field that we can start to address. First, the inclusion criteria may result in a relatively restricted sample, as participants are required to have no concurrent SUD (other than nicotine or cannabis) and no major co-occurring psychiatric disorders. Concurrent substance use is common in people with OUD, with estimates suggesting that more than 25% of individuals have a co-occurring alcohol and/or stimulant use disorder (Jones & McCance-Katz, 2019; Mahoney et al., 2021). These SUDs present concerns regarding safety as well as scientific validity, necessitating limiting their occurrence in this protocol. Similarly, co-occurring psychiatric disorders, such as depression, anxiety and post-traumatic stress disorder (PTSD), are highly prevalent (Jegede, Rhee, Stefanovics, Zhou, & Rosenheck, 2022; Santo Jr et al., 2024). However, it remains an open question whether TMS protocols targeting OUD-related outcomes may convey broader therapeutic benefits. Notably, given that TMS is already cleared for depression (Cohen et al., 2022) and has demonstrated efficacy in PTSD (Harris & Reece, 2021), this study will also collect data on these symptoms to help clarify potential secondary therapeutic benefits. Importantly, recruitment through outpatient clinics that provide buprenorphine treatment will allow for direct collaboration with clinicians to identify appropriate candidates, and inclusion of two study sites will help to broaden the available sample of participants. Second, we will use a novel dosing schedule, with twice-weekly TMS sessions delivered on a single day. While this approach may improve feasibility, the existing literature supports treatment efficacy based on the number of sessions, rather than pulse count alone (Hutton et al., 2023; Schulze et al., 2018; Teng et al., 2017; Yu et al., 2024). As such, the proposed schedule should be considered exploratory, and its effects in this population remain to be established. Third, the optional neuroimaging component raises additional questions about the feasibility of integrating neural biomarkers into clinical protocols of TMS for OUD. While optional participation in this component may reduce participation in this aspect of the study and limit the completeness of mechanistic analyses, the close proximity of neuroimaging facilities may reduce participant burden and facilitate greater participation in imaging procedures.

## Conclusion

If successful, this study will identify a safe and potentially effective adjunctive intervention for reducing craving and return to use in individuals with OUD that is feasible and ready to be implemented in TMS clinics. It will also provide insight into the neural mechanisms underlying changes in craving associated with TMS treatment. By targeting craving - a key contributor to continued opioid use - this approach may help improve clinical outcomes and address limitations of existing treatments for a condition associated with substantial morbidity and mortality.

## Data Availability

No datasets were generated or analysed during the current study.

## Acknowledgements

Author Contributions

All authors read and approved the submitted version of the manuscript. K.B. and H.B.W. drafted the manuscript. All authors made substantial contributions to the design of the work and revised the manuscript.

